# Use of a humanized anti-CD6 monoclonal antibody (itolizumab) in elderly patients with moderate COVID-19

**DOI:** 10.1101/2020.07.24.20153833

**Authors:** Yayquier Díaz, Mayra Ramos-Suzarte, Yordanis Martín, Néstor Antonio Calderón, William Santiago, Orlando Viñet, O Yulieski La, Jorge Pérez, Augusto Oyarzábal, Yoan Pérez, Geidy Lorenzo, Meylan Cepeda, Danay Saavedra, Zaima Mazorra, Daymys Estevez, Patricia Lorenzo-Luaces, Carmen Valenzuela, Armando Caballero, Kalet Leon, Tania Crombet, Carlos Jorge Hidalgo

**Affiliations:** Manuel Fajardo Universitary Hospital, Santa Clara, Villa Clara, Cuba; Center of Molecular Immunology, Playa, La Habana, Cuba; Arnaldo Milian Castro Universitary Hospital. Santa Clara, Villa Clara. Cuba

**Keywords:** Itolizumab, elderly, COVID-19, SARS-CoV-2, immunomodulatory drugs

## Abstract

**Introduction:** The Severe Acute Respiratory Syndrome Coronavirus 2 (SARS-CoV-2) has caused a recent outbreak of Coronavirus Disease (COVID-19). In Cuba, the first case of COVID-19 was reported on March 11. Elderly with multiple comorbidities are particularly susceptible to adverse clinical outcomes in the course of SARS CoV-2 infection. During the outbreak, a local transmission event took place in a nursing home in Villa Clara province, Cuba, in which nineteen elderly residents were positive for SARS-CoV-2.

**Methods:** Based on the increased susceptibility to viral-induced cytokine release syndrome inducing respiratory and systemic complications in this population, the patients were included in an expanded access clinical trial to receive itolizumab, an anti-CD6 monoclonal antibody.

**Results:** All the patients had underlying medical conditions. The product was well tolerated. After the first dose, the course of the disease was favorable and 18 out of 19 (94.7%) patients were discharged clinically recovered with negative RT-PCR at 13 days (median). One dose of itolizumab, circulating IL-6 decreased in the first 24-48 hours in patients with high baseline values, whereas in patients with low levels, this concentration remained over low values. To preliminary assess the effect of itolizumab, a control group was selected among the Cuban COVID-19 patients, which did not receive immunomodulatory therapy. Control subjects were well-matched regarding age, comorbidities and severity of the disease. Every three moderately ill patients treated with itolizumab, one admission in intensive care unit (ICU) was prevented.

**Discussion/Conclusion:** Itolizumab was well tolerated. Its effect is associated with a reduction and controlling IL-6 serum levels. Moreover, treated patients had a favorable clinical outcome, considering their poor prognosis. This treatment is associated significantly with a decrease the risk to be admitted in ICU and reduced 10 times the risk of death. This study corroborates that the timely use of itolizumab, in combination with other antiviral and anticoagulant therapies, is associated with a reduction the COVID-19 disease worsening and mortality. The humanized antibody itolizumab emerges as a therapeutic alternative for patients with COVID-19 and suggests its possible use in patients with cytokine release syndrome from other pathologies.

## Introduction

From December 2019, the first cases of coronavirus disease 2019 (COVID 19) pneumonia began to be registered in the Chinese city of Wuhan. A month later, the World Health Organization (WHO) confirms that the cause of this pneumonia is the severe acute respiratory syndrome coronavirus 2 (SARS-CoV-2). To date (June 23, 2020), 185 countries are reported with cases of COVID-19 and it amounts to 9 million 063,264 confirmed cases, 471 681 deaths for a fatality of 5.20%. In the Americas region, 4 million 512,775 confirmed cases are reported (49.79% of the total reported cases in the world) with a fatality of 5.02%. The first positive case in Cuba was reported on March 11 and to date, 2319 positive cases have accumulated, with 85 deaths reported (June 25, 2020) [1].

COVID-19 can range from asymptomatic to critical disease. Many patients, mostly younger, are asymptomatic or minimally symptomatic. Elderly with pre-existing comorbidities have been at higher risk of severe disease and fatal outcome [2]. Changes occurring in the immune system with increasing age, termed immunosenescence and the state of chronic, low-grade inflammation known as inflammaging, characterized the immune system in the elderly. Both processes suggest the origin of most of the comorbidities of the older adults and its susceptibility to suffer cancer, chronic inflammatory diseases and new infections [3].

SARS-CoV-2 infection is reported to trigger a pro-inflammatory response characterized by high levels of cytokines, including IL-6 and overexpression of other growth factors such as fibroblast growth factor (FGF), the granulocyte macrophage colony stimulating factor (GM-CSF), tumor necrosis factor (TNFα) and vascular endothelial growth factor (VEGF) among others. In some vulnerable patients, the disease might worsen up to admission into the intensive care unit (ICU), if appropriate therapy is not used. Patients at this stage show high levels of pro-inflammatory cytokines. Particularly, many authors have found that IL-6 levels are directly correlated with increased mortality and inversely with lymphocyte count. Based on these findings it has been suggested that cytokine release syndrome may hinder the adaptive immune response against SARS-CoV-2 infection [4, 5].

High number of COVID-19 associated deaths has been seen in the nursing home sector. It is suggested that elderly patients residing in long-term care facilities (LTCFs) could be more vulnerable to SARS-CoV-2 infection becoming a population with higher risk for COVID-19-associated morbidity and mortality [6].

The Center of Molecular Immunology developed the humanized monoclonal antibody (MAb) itolizumab (CIMAREG) capable of binding to a region in the distal membrane domain of human CD6 (domain 1) [7]. The *in vitro* characterization of its mechanism of action revealed that it reduces the expression of intracellular proteins involved in activation and inhibits the proliferation of T cells. The effect is associated with the reduction of activation signals and the production of pro-inflammatory cytokines (interferon-γ (IFN-γ), interleukin (IL)-6 and tumor necrosis factor alpha (TNF-α)) [8]. Studies in blood and tissue samples from patients with severe psoriasis treated with this MAb, showed that the treatment reduced the proliferation capacity of T cells and the number of T cells producing IFN-γ. In addition, the reduction of serum IL-6 and IFN-γ levels was observed [9, 10].

A local transmission event occurred in a Cuban’s nursing home, in which several patients were positive for SARS-CoV-2. Nineteen of these patients were admitted to a hospital at moderate stage of the disease. All the patients were older than 64 years and the majority of them suffered hypertension and/or other comorbidities, which are risk factors associated with severity and fatal COVID-19 outcome. Based on the immunoregulatory action of itolizumab and the safety profile verified in previous severe COVID-19 patients, its use was proposed in the moderately ill elderly patients from the nursing home.

In this study we characterized the clinical features and outcomes of these nineteen elderly patients treated with itolizumab combined with anti-viral therapies.

## Materials and Methods

### Patients

Nineteen patients confirmed as SARS-CoV-2 positive by real-time transcriptase polymerase chain reaction (RT-PCR), admitted from the Cuban’s nursing home, were enrolled in an open, multicenter expanded access clinical trial (RPCEC00000311(VICTORIA)) to be treated with the humanized MAb itolizumab. The Ethics Committee for Clinical Research in the hospital and Cuban Regulatory Agency (CECMED) approved the trial. The study complied with the Good Clinical Practices and the precepts established in the Declaration of the Helsinki World Medical Association. All patients met the inclusion criteria described in the protocol (http://rpcec.sld.cu/trials/RPCEC00000311-Eg.) and signed the informed consent.

### Treatment

Patients received standard treatments (lopinavir/ritonavir (kaletra), chloroquine, prophylactic antibiotics, interferon alpha (α) 2B and low-molecular-weight heparin (LMWH)) included in the institutional protocol for COVID-19 [11].

In addition, patients received a first intravenous dose of 200 mg of a humanized Mab itolizumab (8 vials). Some patients received a second dose (200 mg), considering their clinical evolution and the physician criteria. Itolizumab-associated adverse events (AE) were reported. Their classification was performed according to the NIH-CTC toxicity criteria, version 5.0.

### Laboratory examination

Laboratory results included blood routine, leucocyte subsets and blood biochemical parameters. The level of inflammatory cytokines was measured using human Quantikine ELISA Kits from R&D Systems (Minneapolis, USA): Human IL-6 Quantikine ELISA Kit (Cat# S6050).

### Statistical Analysis

Continuous variables were expressed as mean and standard deviations or median and interquartile ranges (depending of the distribution of each variable). Wilcoxon rank sum test were applied to continuous variables, and chi-square or Fisher’s exact test were used for categorical variables. Type 1 error of 0.05 was used. To preliminary assess the effect of itolizumab, a control group was selected among the Cuban COVID-19 patients, that did not receive immunomodulatory therapy. Controls were treated with lopinavir/ritonavir, chloroquine, interferon α2B and LMWH. Control subjects were well-matched regarding age, comorbidities and severity of the disease. Odds ratio for disease progression and mortality was estimated for itolizumab vs control. The number of patients needed to treat (NNT) to prevent an additional poor outcome and the absolute risk reduction were also calculated. The analysis was performed using SPSS 25.0 software.

## Results

### Baseline characteristics of COVID-19 elderly patients

The median age was 79 years (64 to 100 years old); 63.2% were female and 68.4% were white skin. All the patients had chronic underlying health conditions. The most frequent comorbidities were hypertension (73.7%), dementia (57.9%), malnutrition (52.6%), cardiac disease (42.1%), diabetes mellitus (31.6%) and chronic obstructive pulmonary disease (COPD) (26.3%). The majority of them suffered more than one comorbidity (Table 1).

**Table 1.** Demography characteristic of elderly COVID-19 patients, treatments and outcomes.

| Case No. | Sex | Co-morbidities | Treatments additional | Outcomes |
| --- | --- | --- | --- | --- |
| 12 | F | Hypertension, Ischemic heart disease, Malnutrition, Dementia | itolizumab (2 doses), IFNα2B, Kaletra, Chloroquine, LMWH, parenteral Vitamins | Alive |
| 13 | M | Hypertension, Chronic Obstructive Pulmonary disease, Dementia | itolizumab (2 doses), IFNα2B, Kaletra, Chloroquine LMWH, parenteral Vitamins | Alive |
| 14 | F | Malnutrition, Dementia | itolizumab (2 doses), IFNα2B, Kaletra, Chloroquine, LMWH, parenteral Vitamins | Alive |
| 15 | F | Hypertension, Diabetes Mellitus, Malnutrition, Dementia | itolizumab (2 doses), Kaletra, Chloroquine, LMWH, parenteral Vitamins, Antibiotics. | Alive |
| 16 | F | Hypertension, Diabetes Mellitus, Chronic Obstructive Pulmonary disease, Asthma, Malnutrition | itolizumab (2 doses), IFNα2B F, Kaletra, Chloroquine, LMWH, parenteral Vitamins | Alive |
| 17 | M | Hypertension, Diabetes Mellitus, Chronic Obstructive Pulmonary disease, Dementia, Smoker, Hypertensive heart disease | itolizumab (2 doses), IFNα2B, Kaletra, Chloroquine, LMWH, parenteral Vitamins | Alive |
| 18 | F | Dementia, Anemia | itolizumab (2 doses), IFNα2B, Kaletra, Chloroquine, LMWH, Antibiotics | Alive |
| 19 | F | Hypertension, Diabetes Mellitus, Ischemic heart disease, Malnutrition | itolizumab (2 doses), Kaletra, Chloroquine, LMWH, Antibiotics | Alive |
| 20 | F | Hypertension, Ischemic heart disease, Heart disease | itolizumab (2 doses), IFNα2B, Kaletra, Chloroquine, Heparin, Antibiotics | Alive |
| 21 | M | Hypertension, Chronic Obstructive Pulmonary disease | itolizumab (2 doses), IFNα2B, Kaletra, Chloroquine, Heparin, Vitamins | Alive |
| 22 | F | Hypertension, Diabetes Mellitus, Ischemic heart disease, Dementia | itolizumab (2 doses), Kaletra, Chloroquine, LMWH, parenteral Vitamins | Alive |
| 24 | F | Hypertension, Ischemic heart disease, Malnutrition, Dementia | itolizumab (2 doses), Kaletra , Chloroquine, LMWH, parenteral Vitamins | Alive |
| 25 | M | Hypertension, Diabetes Mellitus, Malnutrition, Dementia, amputated | itolizumab (2 doses), IFNα2B, Kaletra, Chloroquine, LMWH, parenteral Vitamins | Alive |
| 26 | F | Hypertension, Ischemic heart disease, Malnutrition, Dementia | itolizumab (2 doses), IFNα2B, Kaletra, Chloroquine, LMWH, parenteral Vitamins | Death |
| 27 | F | Hypertension, Ischemic heart disease, Malnutrition, Dementia | itolizumab (2 doses), Kaletra, Chloroquine, LMWH, parenteral Vitamins | Alive |
| 32 | F | Hypertension, Ischemic heart disease, Chronic Obstructive Pulmonary disease, Malnutrition | itolizumab (2 doses), IFNα2B, Kaletra | Alive |
| 40 | M | None | itolizumab (1 dose), IFNα2B, Kaletra, Chloroquine, Heparin, Erythropoietin | Alive |
| 43 | M | Obesity, Smoker, Alcoholism, Post traumatic paraplegia | itolizumab (2 doses), Kaletra, Chloroquine, Heparin, Erythropoietin | Alive |
| 44 | M | None | itolizumab (1 dose), IFNα2B, Kaletra, Chloroquine, Antibiotics, Erythropoietin, Steroids, Omeprazole | Alive |

IFNα2B: Interferon alpha 2B; LMWH: low-molecular-weight heparin; N.A: non available

Upon admission, 42.1% of these elderly patients required oxygen therapy due to respiratory function worsening. In addition, 52.6% had fever, 47.3% had cough and 42.1% showed signs and symptoms of dehydration. Other symptoms included dyspnea (36.8%) and diarrhea (26.3%). Regarding the severity of the disease, they were classified as moderately ill patients.

Concerning hematological and biochemical parameters, 16.7% of patients had leukocyte levels below the normal range. Lymphocytes were below the normal range in 29.4% and 5.6% had low counts of platelets. Aspartate aminotransferase (ASAT) was above the normal values in 13.3% and D-dimer was increased in 84.6% of the patients.

### Treatment with an anti-CD6 monoclonal antibody (itolizumab)

Immediately upon confirmation of the presence of SARS-CoV-2, the patients received the established institutional COVID-19 treatment protocol [11]. All the patients received lopinavir/ritonavir (kaletra), 94.7% had chloroquine, 68.4% received interferon α2B and 89.5% received LMWH. Additionally, 63.2% got vitamins and 31.6% antibiotics. Three patients (15.8%) were treated with recombinant human erythropoietin at a cytoprotective dose (Table 1).

All the patients received one dose of the antibody, while 89.5% received two. The median time between the onset of symptoms and itolizumab was 1 day. The time between doses, ranged from 1 to 7 days, with an overall median of two days. In this study, 94.7% of the patients were discharged, with a median hospital stay of 13 days, ranging from 3 to 40 days. All of them were negative for SARS-COV2 at the moment of discharge. Only one death-event took place, in a patient with several comorbidities (hypertension, coronary artery disease, dementia and malnutrition). In it, daily electrocardiograms showed a left bundle branch block without sings of acute ischemia and death was by pulmonary thromboembolism. Only one AE related to the administration of the itolizumab was reported. The AE consisted on chills and occurred immediately after the first administration, lasting for minutes. The AE was mild intensity and did not imply changes in the administration of the antibody.

Vital signs were evaluated after itolizumab administration during the hospital stay. After 7 days of itolizumab treatment, only two patients required oxygen therapy (10.5%) and this difference had statistically significant compared to baseline requirements (42.1% vs 10.5%, p=0.031, χ^2^ (McNemar) test).

IL-6 serum levels were measured before and 24-48 hours after the treatment in twelve elderly patients. The median of pretreatment concentration was 23.9 pg/mL and the median of IL-6 levels 48 hours after the treatment was 25.9 pg/mL. Therefore, no significant changes were detected. In a previous work, the application of receiver operating characteristic (ROC) curve selected the cut-off of IL-6 levels on 28.3 pg/mL, to stablish the association between baseline IL-6 concentration and severity of illness (unpublished data). When analyzing the IL-6 serum values of elderly patients regarding the selected cut-off, five patients had baseline levels above 28.3 pg/mL. Four out of five patients reduced the cytokine concentrations 24-48 hours after the itolizumab. In one patient, the levels remained similar (shown in Fig. 1).

**Fig. 1.**
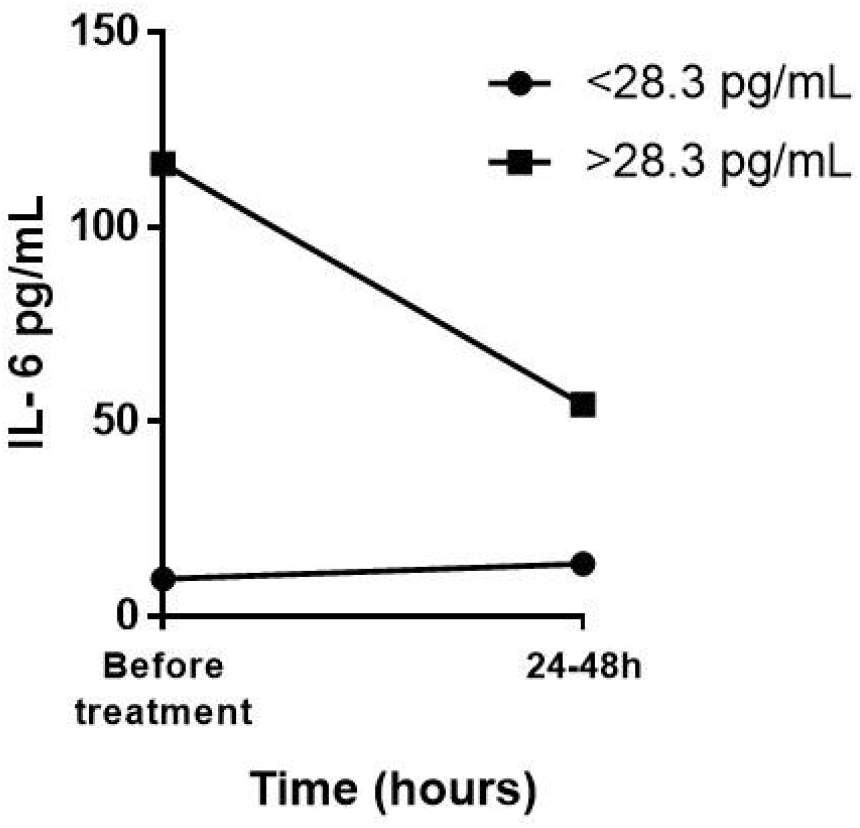
Median of IL-6 concentration in the serum of COVID-19 patients before and 24-48 hours after itolizumab treatment. The patients were divided according to the pre-stablished cut-off of IL-6 levels (28.3 pg/mL).

Among the seven patients with baseline levels below the selected cut-off, in six patients the circulating IL-6 did not increase over 28.3 pg/mL 24-48 hours after the administration. Only in one patient, the IL-6 concentration increased (shown in Fig. 1). The magnitude of change of IL-6 among the patients with concentrations above the cutoff has a median of reduction of 48.6 pg/mL. The median of change in IL-6 concentration among the patients with baseline levels below 28.3 pg/mL, was 2.2 pg/mL.

### Clinical outcome of itolizumab-treated patients

A comparison of the clinical evolution between the cohort from the nursing home (n=19) and a group of Cuban COVID-19 patients with similar baseline conditions (control group), was performed. Control patients were defined as COVID-19-confirmed patients, reported by the Cuban Ministry of Public Health. Among them, patients with similar baseline conditions to our cohort: at least one comorbidity (hypertension, diabetes mellitus, cardiac disease, cancer, chronic kidney disease, obesity, malnutrition or COPD), 64 years or older and who were not included in other COVID-19 clinical trials, were selected (n=53 patients). Control patients received the rest of the Cuban protocol drugs described above. The groups were homogeneous in terms of demographic characteristics and significant comorbidities, except for malnutrition, which occurs only in the itolizumab treated group (p=0.000, test χ^2^).

Analyzing control and itolizumab-treated patients with two or more comorbidities, a significant dependence between itolizumab use and admission in intensive care unit (ICU) or mortality, was detected. Every three moderately ill patients treated with itolizumab, one admission in ICU was prevented (p=0.042, test χ^2^, NNT: 3.12). Additionally, treatment with the Mab is associated with a reduction the risk of death 10 times as compared with the control group (1/0.10=10). Every three moderately ill patients treated with itolizumab, one death was prevented (p=0.020, test χ^2^, NNT: 2.93) (shown in Fig. 2).

**Fig. 2.**
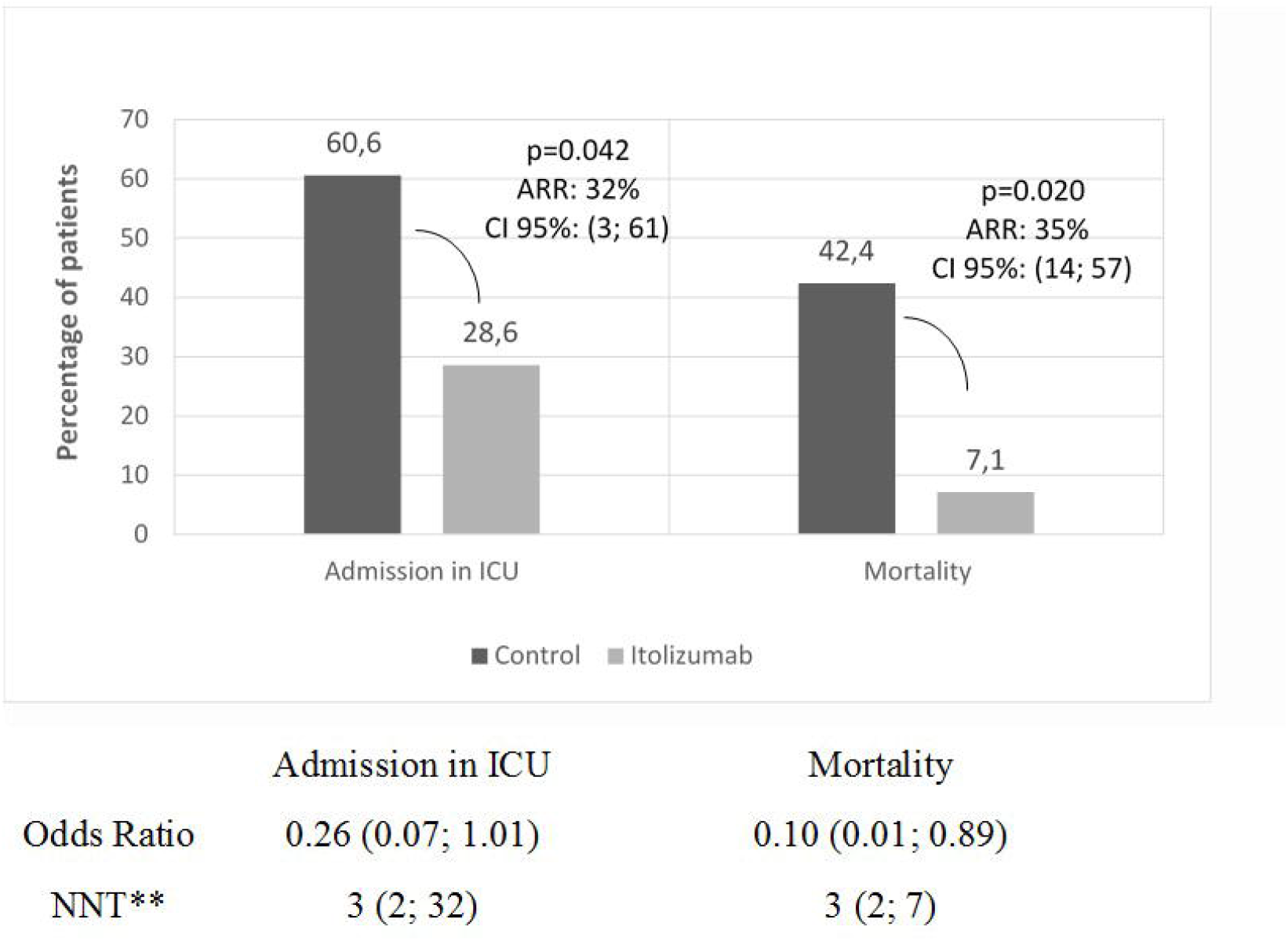
Clinical outcome of itolizumab-treated and control patients. Frequency of patients admitted in ICU and mortality. Odds ratio for disease progression and mortality, NNT to prevent an additional poor outcome (admission in the ICU, death) and the ARR were calculated for itolizumab and control patients. Difference were considered significant if p< 0.05. *ARR: Absolute Risk Reduction (%);** NNT: Number needed to treat. ICU: intensive care unit. CI: confident interval.

## Discussion

Elderly patients are particularly susceptible to adverse clinical outcomes in the course of SARS CoV-2 infection. Older adults with age-related diseases are more susceptible to viral-induced cytokine storm resulting in respiratory failure, multisystem damage and fatal outcome [12].

COVID-19, caused by the new coronavirus SARS-CoV-2, become a global health emergency according to the WHO since March 11, 2020, has led the international scientific and medical community to search for adequate therapies that would allow controlling it, avoiding mortality and the damages associated with it [12].

In April 2020, an episode of massive SARS CoV-2 transmission occurred in the Nursing Home in Villa Clara province, Cuba. Nineteen residents were infected by a visitor, who turned out to be positive at SARS CoV-2 days after having made a visit to that facility. Residents and health care personnel at long-term care facilities are at risk for COVID-19 transmission and residents are particularly at risk of severe outcomes because they are predominantly at advanced ages and have commonly underlying medical conditions [6].

After the appearance of COVID 19-related symptoms, all the elderly were admitted into the Hospital. They immediately received the treatment-protocol established in the country for this disease [11].

All the patients in our cohort had underlying medical conditions. The most frequent were hypertension, dementia, malnutrition, cardiac disease, diabetes mellitus and COPD. The median age was 79 years old. People of any age with certain comorbidities such as hypertension, COPD, serious heart conditions, diabetes mellitus and immunocompromised state, especially elderly, are at increased risk for severe illness from COVID-19 [13]. Fever, cough, dyspnea, signs of dehydration and diarrhea were the most frequent symptoms on admission. They were classified as moderately ill patients. Assessment and treatment is challenging for elderly patients diagnosed with COVID-19. An individualized approach should be offered to older adults properly analyzing the beneficial effects and the risks of therapeutic decisions [12].

Based on the increased susceptibility to viral-induced cytokine storm inducing respiratory and systemic complications in this type of population [12], the patients were included in an expanded access clinical trial to receive itolizumab, an anti-CD6 monoclonal antibody. Itolizumab is a first-in class antagonistic Mab that selectively targets the CD6-ALCAM pathway, reducing the activation, proliferation and differentiation of T cells into pathogenic effector T cells and leading to a decrease of pro-inflammatory cytokine production [8].

The product was well tolerated since only one mild adverse reaction was reported after the first infusion in one subject. In general, after the first dose, the course of the disease was favorable, only two patients required oxygen therapy after the treatment and 94.7% of the patients were discharged with negative RT-PCR results at 14 days. This outcome suggests that although itolizumab is an immunomodulatory drug, it did not interfere with the clearance of SARS-CoV-2 virus.

Inflammation arises as a critical process in SARS-CoV-2-infected patients. While a strong immune response may contain the infection, previous studies have reported worse outcomes related to the presence of an excessive number of cytokines and inflammatory soluble factors [14]. Itolizumab is associated with a reduction or stabilized IL-6 levels in these elderly patients in the first 24-48 hours of administration. In four out of five patients with IL-6 baseline concentrations above the pre-stablish cutoff for severity (28.3 pg/mL) (unpublished data), the levels decreased after Mab administration. In one patient, the levels remained similar. Regarding patients with IL-6 baseline levels below 28.3 pg/mL, only one increased the IL-6 values; the rest remained under this cut-off 24-48 hours after the administration. Therefore, these findings suggest that the reduction of serum IL-6 levels in patients with high baseline concentrations, and its stabilization over low values is an effect associated of this monoclonal antibody.

Modulation of the severe inflammatory state in patients with COVID-19, consist in a relevant strategy to limit the severity of pulmonary and systemic complications. This approach could successfully reduce the needs for intensive care support and mechanical ventilation, and eventually decrease mortality [15]. When comparing our cohort of COVID-19 itolizumab-treated patients with a control group of Cuban COVID-19 patients, with similar characteristics (age, comorbidities and similar treatment except for itolizumab or another immunomodulatory agent), itolizumab emerged as therapy associated on preventing the severity of the disease. This treatment is associated significantly with a decrease the risk to be admitted in ICU and reduced 10 times the risk of death.

This study has some limitations regarding the short kinetic of IL-6 levels and the limited follow up of the patients, long-term outcomes of them are unknown. Nevertheless, it shows the effect of itolizumab associated with a reduction and controlling IL-6 serum levels. Moreover, itolizumab treated patients had a favorable clinical outcome, considering their poor prognosis associated with old age and comorbidities, including nutrition deficit.

In conclusion, this study corroborates that the timely use of itolizumab, in combination with other antiviral and anticoagulant therapies, is associated with a reduction the COVID-19 disease worsening and mortality. The humanized antibody itolizumab emerges as a therapeutic alternative for patients with COVID-19 and suggests its possible use in patients with cytokine release syndrome from other pathologies.

## Data Availability

All data referred to in the manuscript are available

## Statements

## Acknowledgement

The authors are extremely thankful to our patients and their relatives and to the research teams from the Manuel Fajardo Hospital and from Center of Molecular Immunology.

## Statement of Ethics

The Ethics Committee approved the clinical trial. The Cuban Regulatory Agency (CECMED) also approved the trial. The study complied with the Good Clinical Practices and the precepts established in the Declaration of the Helsinki World Medical Association. All patients signed the informed consent.

## Author Contributions

(1, 3, 4) Yayquier Díaz, Mayra Ramos-Suzarte, Carlos Jorge Hidalgo, Armando Caballero, Kalet Leon, Tania Crombet, Yordanis Martín, Néstor Antonio Calderón, William Santiago, Orlando Viñet, Yulieski La O, Jorge Pérez, Augusto Oyarzábal, Yoan Pérez, Geidy Lorenzo, Meylan Cepeda Portales, Danay Saavedra, Zaima Mazorra, Daymys Estevez, Patricia Lorenzo-Luaces, Carmen Valenzuela

(2) Mayra Ramos-Suzarte, Tania Crombet, Danay Saavedra, Zaima Mazorra, Carmen Valenzuela, Yayquier Díaz, Carlos Jorge Hidalgo.

